# Symptoms of common mental disorders and adherence to antiretroviral therapy among adults living with HIV in rural Zimbabwe: a cross-sectional study

**DOI:** 10.1101/2021.05.22.21257636

**Authors:** Andreas D Haas, Cordelia Kunzekwenyika, Stefanie Hossmann, Josphat Manzero, Janneke H van Dijk, Ronald Manhibi, Ruth Verhey, Andreas Limacher, Per von Groote, Ethel Manda, Michael Hobbins, Dixon Chibanda, Matthias Egger, for IeDEA Southern Africa

**Author notes:** Correspondence to: Andreas D Haas, Institute of Social & Preventive Medicine, University of Bern, Mittelstrasse 43, CH-3012 Bern, Switzerland.

## Abstract

**Objectives:** To examine the proportion of people living with HIV who screen positive for common mental disorders (CMD) and the associations between CMD and self-reported adherence to antiretroviral therapy (ART).

**Setting:** Sixteen government-funded health facilities in the rural Bikita district of Zimbabwe.

**Design:** Cross-sectional survey.

**Participants:** HIV-positive non-pregnant adults, aged 18 years or older, who lived in Bikita district and had received ART for at least six months.

**Outcome measures:** The primary outcome was the proportion of participants screening positive for CMD defined as a Shona Symptoms Questionnaire (SSQ-14) score of 9 or greater. Secondary outcomes were the proportion of participants reporting suicidal ideation, perceptual symptoms, and suboptimal ART adherence and adjusted prevalence ratios (aPR) for factors associated with CMD, suicidal ideation, perceptual symptoms, and suboptimal ART adherence.

**Results:** Out of 3,480 adults, 18.8% (95% confidence interval [CI] 14.8-23.7) screened positive for CMD, 2.7% (95% CI 1.5-4.7) reported suicidal ideations, and 1.5% (95% 0.9-2.6) reported perceptual symptoms. Positive CMD screens were more common in women (adjusted prevalence ratio [aPR] 1.67, 95% CI 1.19-2.35) than in men and were more common in adults aged 40-49 years (aPR 1.47 95% CI 1.16-1.85) or aged 50-59 years (20.3%; aPR 1.51 95% CI 1.05-2.17) than in those 60 years or older. Positive CMD screen was associated with suboptimal adherence (aPR 1.53; 95% CI 1.37-1.70).

**Conclusions:** A substantial proportion of people living with HIV in rural Zimbabwe are affected by CMD. There is a need to integrate mental health services and HIV programs in rural Zimbabwe.

**Strengths and limitations of this study:**

- Inclusion of a large sample of people living with HIV recruited at 16 government-funded primary and secondary care facilities in a rural district of Zimbabwe.
- Use of a locally developed screening tool that showed good psychometric properties for detecting common mental disorders in Zimbabwe in HIV-positive urban populations.
- The screening tool was not validated for the rural setting, and the cutoff score was selected based on data from the urban setting.
- Adherence to antiretroviral therapy was self-reported.

## Background

In 2019, approximately 1.4 million people were living with HIV in Zimbabwe, of whom more than 1.1 million were receiving antiretroviral therapy (ART) [1]. Widespread access to ART has dramatically improved the life expectancy of people living with HIV [2]. However, the long-term effectiveness of ART depends on lifelong retention in HIV care and strict medication adherence [3,4].

Common mental disorders (CMD), which include depression and anxiety disorders are highly prevalent among people living with HIV. In sub-Saharan Africa, the estimated prevalence of major depression in people living with HIV is 15.3%, and of depressive symptoms 27.0% [5]. The median prevalence of anxiety disorders in people living with HIV in developing countries is estimated at 22.8% [6]. The prevalence of depression and anxiety disorders is higher in women than in men [7,8]. In Zimbabwe’s capital Harare, over 50% of people living with HIV attending a primary care facility met diagnostic criteria for either depression or anxiety, and 65% screened positive for CMD [9,10]. In Zimbabwe, most people living with HIV receiving ART reside in rural areas [11], and the CMD prevalence in this population is unknown.

The co-occurrence of mental disorders and HIV poses significant challenges in managing and treating HIV. Mental disorders are associated with poor HIV treatment outcomes, including low adherence, lack of viral load suppression, loss to follow-up and mortality [12–16]. Early detection and effective management of mental disorders among people living with HIV may improve the quality of life of affected individuals, ART adherence and viral load suppression [17,18], thus reducing the incidence of HIV-associated complications, preventing drug resistance and HIV transmission. Despite these benefits, there is a large ‘treatment gap’ in mental health care among people living with HIV in low- and middle-income countries: most people affected by mental illness do not receive appropriate treatment [19,20].

The Friendship Bench intervention is a culturally adapted evidence-based psychological intervention developed to close the treatment gap for CMD in Zimbabwe [21]. The Friendship Bench team trains community health workers to identify people with CMD symptoms and deliver a brief intervention consisting of six sessions of problem-solving therapy and optional group support [21]. We are conducting a cluster-randomised trial to assess the effectiveness of the Friendship Bench intervention in improving ART outcomes and symptoms of CMD in people living with HIV in rural Zimbabwe. During recruitment, we screened over 3,500 ART patients for CMD and poor adherence. In this paper, we report the prevalence of positive CMD screening tests and associations between positive CMD screens and self-reported adherence among people living with HIV in rural Zimbabwe.

## Methods

We conducted a cross-sectional study at 16 health facilities in Bikita district to assess the eligibility of individuals for a cluster-randomised trial (FB-ART) on the effect of the Friendship Bench intervention [22] on ART outcomes and symptoms of CMD in people living with HIV. Bikita is a rural district of the Masvingo Province about 300 km south of Harare. We registered the trial with ClinicalTrials.gov (NCT03704805).

Between October 5, 2018, and December 19, 2019, we offered CMD screening at 16 government-funded health facilities in rural Zimbabwe. HIV-positive non-pregnant adults aged 18 years or older who lived in Bikita district and had received ART for at least six months were eligible. Trained research assistants offered screening for CMD using the Shona Symptoms Questionnaire (SSQ-14) [9] and assessed adherence using a question from the AIDS indicator survey [23]. The SSQ-14 is a locally developed CMD screening tool [9]. The tool assesses if individuals had experienced common mental health symptoms, including sleep disturbance, suicidal ideations, tearfulness, perceptual symptoms, and impairment of functioning in the past seven days. Each of the 14 symptoms is scored dichotomously (symptom present or absent) [24]. The tool showed good psychometric properties for detecting CMD in HIV-positive and HIV-negative urban populations in Zimbabwe [9]. An SSQ-14 score of ≥9 had a sensitivity of 88% and a specificity of 76% for depression or general anxiety in HIV-positive adults in Harare [9]. The tool had a high internal consistency in the validation study (Cronbach’s α=0.74) and in our study (Cronbach’s α=0.82). Adherence was assessed based on self-report using the following question: “In the last 30 days, how many days have you missed taking any of your ARV [antiretroviral] pills?” [23].

We defined SSQ-14 scores of 9 or greater as positive CMD screen [9]. Participants who reported that they “felt like committing suicide” in the past seven days (SSQ-14 item 11) screened positive for suicidal ideation. Participants who reported that they “saw or heard things which others could not see or hear” in the past seven days (SSQ-14 item 5) screened positive for perceptual symptoms. Participants who indicated that they had missed taking one or more ARV pills in the last 30 days were classified as having suboptimal adherence. Those reporting not having missed any ARV pills had optimal adherence. We categorised age into the following groups: 18-29 years, 30-39 years, 40-49 years, 50-59 years, and 60 years or older.

Individuals who participated in SSQ screening were eligible for this analysis. Participants who did not respond to all 14 SSQ items were excluded. We calculated the prevalence of non-adherence and positive screening outcomes with logit-transformed 95% confidence intervals (CI) that adjusted for intragroup correlation at health facilities. We estimated adjusted prevalence ratios (aPRs) for factors associated with positive screening for CMD, suicidal ideation, and perceptual symptoms using mixed-effects Poisson regression models with robust standard errors [25]. Models were adjusted for sex, age, and clustering of data at facility-level using a random intercept for study facilities. We used the same models to calculate unadjusted and aPRs for factors associated with suboptimal adherence. Statistical analysis was done using Stata (Version 16, Stata Corporation, College Station, TX, USA).

The study protocol was approved by the ethics committees of the Medical Research Council of Zimbabwe (MRCZ), the Research Council of Zimbabwe (RCZ), and the Canton of Bern, Switzerland. Individuals provided verbal consent for eligibility screening and collection and analysis of screening data. Research assistants referred individuals screening positive for CMD with suicidal ideation or perceptual symptoms to the nurse in charge for further assessment and care. Individuals who screened positive for CMD and provided written informed consent were included in the trial and were offered CMD treatment as part of the FB-ART trial.

### Patient and Public Involvement

Patients with psychiatric morbidity were involved in ethnographic and qualitative research, which informed the development of the SSQ-14 [24]. The results of this study were shared with the provincial medical director and the district medical officer.

## Results

Research assistants assessed the eligibility for CMD screening of 3,707 individuals: 3,543 (95.6%) of eligible individuals participated in CMD screens. Pregnancy, residency outside of Bikita district, and age below 18 years were the most common reasons for ineligibility for screening. Out of 3,543 individuals screened for CMD, 63 (1.7%) did not respond to all SSQ-14 items and were excluded. The remaining 3,480 individuals were included in the analysis. The median age of the study population was 45 years (IQR 38–53). Three-quarters of the participants (74.9%, n=2,608) were women.

Table 1 shows the prevalence and aPRs for positive screening for CMD, suicidal ideation, and perceptual symptoms. Out of 3,480 adults, 18.8% (95% CI 14.8-23.7, n=655) screened positive for CMD, 2.7% (95% CI 1.5-4.7, n=93) reported suicidal ideation, and 1.5% (95% CI 0.9-2.6, n=52) reported perceptual symptoms. Positive CMD screens were more common in women (21.0%; aPR 1.67, 95% CI 1.19-2.35) than in men (12.4%) and were more common in adults aged 40-49 years (20.6%; aPR 1.47 95% CI 1.16-1.85) or aged 50-59 years (20.3%; aPR 1.51 95% CI 1.05-2.17) than in those 60 years or older (12.6%). Suicidal ideations were more common in adults 18-29 years (4.7%, 95% CI 0.2-9.2) and in adults aged 30-39 years (4.0%, 95% CI 1.1-7.0) than in older adults, but the statistical uncertainty around these estimates was large.

**Table 1:** Prevalence of symptoms of common mental disorders, suicidal ideation, and perceptual disorders among people living with HIV in rural Zimbabwe.

| Positive screens |  |  |  |  |  |  |  |  |  |  |
| --- | --- | --- | --- | --- | --- | --- | --- | --- | --- | --- |
|  | Common mental disorders<sup>a</sup> |  |  |  | Suicidal ideation<sup>b</sup> |  |  | Perceptual symptoms<sup>c</sup> |  |  |
|  | Total screened<br>N | N | Prevalence<sup>d</sup><br>(95% CI) | Adjusted prevalence ratio<sup>e</sup><br>(95% CI) | N | Prevalence<sup>d</sup><br>(95% CI) | Adjusted prevalence ratio<sup>e</sup><br>(95% CI) | N | Prevalence<sup>d</sup><br>(95% CI) | Adjusted prevalence ratio<sup>e</sup><br>(95% CI) |
|  | 3,480 | 655 | 18.8%<br>(14.8-23.7) |  | 93 | 2.7%<br>(1.5-4.7) |  | 52 | 1.5%<br>(0.9-2.6) |  |
| Sex |  |  |  |  |  |  |  |  |  |  |
| Female | 2,654 | 547 | 21.0%<br>(16.5-26.2) | 1.67<br>(1.19-2.35) | 78 | 3.0%<br>(1.2-4.7) | 1.47<br>(0.71-3.03) | 43 | 1.6%<br>(0.7-2.6) | 1.47<br>(0.73-2.96) |
| Male | 886 | 108 | 12.4<br>(6.9-17.9) | 1.00 | 15 | 1.7%<br>(0.2-3.3) | 1.00 | 9 | 1.0%<br>(0.1-2.0) | 1.00 |
| Age in years |  |  |  |  |  |  |  |  |  |  |
| 18-29 | 280 | 49 | 17.8%<br>(10.3-25.3) | 1.19<br>(0.74-1.92) | 13 | 4.7%<br>(0.2-9.2) | 2.21<br>(0.86-5.66) | 4 | 1.5%<br>(0.0-2.9) | 2.47<br>(0.69-8.78) |
| 30-39 | 761 | 138 | 18.5%<br>(13.6-23.4) | 1.28<br>(0.88-1.87) | 30 | 4.0%<br>(1.1-7.0) | 1.65<br>(0.73-3.71) | 12 | 1.6%<br>(0.0-3.3) | 2.54<br>(0.44-14.57) |
| 40-49 | 1,292 | 262 | 20.6%<br>(14.9-26.3) | 1.47<br>(1.16-1.85) | 28 | 2.2%<br>(0.6-3.8) | 0.87<br>(0.51-1.49) | 23 | 1.8%<br>(0.5-3.2) | 2.73<br>(0.87-8.54) |
| 50-59 | 746 | 149 | 20.3%<br>(15.2-25.5) | 1.51<br>(1.05-2.17) | 12 | 1.6%<br>(0.6-2.7) | 0.73<br>(0.43-1.24) | 10 | 1.4%<br>(0.4-2.4) | 2.13<br>(0.63-7.20) |
| 60+ | 461 | 57 | 12.6%<br>(6.9-18.3) | 1.00 | 10 | 2.2%<br>(0.7-3.7) | 1.00 | 3 | 0.7%<br>(0.0-1.4) | 1.00 |

Data are numbers of individuals, the prevalence of positive screens, and adjusted prevalence ratios. 95% confidence intervals are shown in parenthesis.
<sup>a</sup> Shona Symptoms Questionnaire (SSQ-14) scores of 9 or greater
<sup>b</sup> Yes to item 11 on the SSQ-14: "At times I felt like committing suicide."
<sup>c</sup> Yes to item 5 on the SSQ-14: "I sometimes saw or heard things which others could not see or hear."
<sup>d</sup> Prevalence of positive screening tests and logit-transformed confidence intervals adjusted for intragroup correlation at health facilities.
<sup>e</sup> Models adjusted for sex, age, and clustering of participants in facilities.
Abbreviations: aPR=adjusted prevalence ratio; CI=confidence interval; SSQ-14= Shona Symptoms Questionnaire.

Out of 3,469 individuals who responded to the adherence question (11, 0.32% did not respond), 2,900 (83.6% 95% CI 80.0-87.2) reported optimal adherence and 569 (16.4% 95% CI 12.8-20.0) reported suboptimal adherence. Suboptimal adherence was more common in individuals screening positive for CMD (aPR 1.53 95% CI 1.37-1.70) than in those screening negative (Table 2). Suboptimal adherence was also more common in men (aPR 1.25 95% CI 1.01-1.53) than in women and adults aged 18-29 years (aPR 1.62 95% CI 1.10-2.38) than in those 60 years or older (Table 2).

**Table 2:** Prevalence ratios for factors associated with suboptimal adherences among people living with HIV in rural Zimbabwe.

|  | Unadjusted<br>prevalence ratio<br>(95% CI)<sup>c</sup> | Adjusted<br>prevalence ratio<sup>d</sup><br>(95% CI) |
| --- | --- | --- |
| CMD screening |  |  |
| Negative<sup>a</sup> | 1.00 | 1.00 |
| Positive<sup>b</sup> | 1.46 (1.29-1.66) | 1.53 (1.37-1.70) |
| Sex |  |  |
| Female | 1.00 | 1.00 |
| Male | 1.17 (0.96-1.44) | 1.25 (1.01-1.53) |
| Age in years |  |  |
| 18-29 | 1.60 (1.10-2.31) | 1.62 (1.10-2.38) |
| 30-39 | 1.29 (0.90-1.85) | 1.31 (0.91-1.89) |
| 40-49 | 1.00 (0.72-1.38) | 0.99 (0.71-1.38) |
| 50-59 | 1.04 (0.72-1.51) | 1.03 (0.71-1.49) |
| 60+ | 1.00 | 1.00 |

Data are unadjusted and adjusted prevalence ratios. 95% confidence intervals are shown in parenthesis.
<sup>a</sup> Shona Symptoms Questionnaire (SSQ-14) scores of smaller than 9.
<sup>b</sup> Shona Symptoms Questionnaire (SSQ-14) scores of 9 or greater.
<sup>c</sup> Models adjusted for clustering of participants in facilities.
<sup>d</sup> Models adjusted for CMD screening, sex, age, and clustering of participants in facilities.
Abbreviations: CI=confidence interval; CMD, common mental disorders ; SSQ-14= Shona Symptoms Questionnaire.

## Discussion

In a rural district of Zimbabwe, about one in five HIV positive adult screened positive for CMD, almost 3% reported suicidal ideation, and 1.5% perceptual symptoms compatible with psychosis. Positive CMD screens were more common in women and middle-aged adults than in men or older adults. Positive CMD screens were associated with suboptimal self-reported ART adherence.

Our results have to be considered in light of two limitations. First, we used a locally developed CMD screening tool that had been validated in urban Zimbabwe, but we did not validate the tool for the rural setting and selected the CMD threshold based on the urban validation study [9]. Second, we used a self-reported adherence measure which might be prone to underreporting of non-adherence. However, there is evidence for the validity of self-reported measures of ART adherence [26]. The strengths of our study include a large sample size and a multicentre design.

In line with the national [10] and international literature [27,28], we found that CMDs were much more common in women than in men. Biological factors, including sex hormones and sex differences in the neuroendocrine response to stress, psychosocial factors such as gender differences in interpersonal orientation, self-esteem, body shaming, and rumination might contribute to the gender gap in CMD [7,8]. In addition to these individual-level factors, gender inequity and higher exposure of women and girls to traumatising life events, including gender-based violence or sexual abuse, may further contribute to the gender gap in CMD [7,10,29].

The prevalence of positive CMD screens observed in our study of a rural HIV positive population was less than a third of the prevalence observed in a study of a similar urban population conducted in Harare in 2013 [9,10]. An important factor that likely contributed to the lower prevalence of CMD in our study compared to earlier studies is the change of national ART guidelines to treat people living with HIV at less advanced stages of HIV disease. In the last decade, the CD4 threshold for ART eligibility was successively raised from <250 cells/µL to immediate ART initiation of all people living with HIV under WHO’s “treat all” guidelines. These changes were reflected in an increase in the median CD4 at ART initiation in low- and middle-income [30,31]. Low CD4 cell count is associated with a higher risk of CMD [32], and the comparably low prevalence of CMD observed in our study might reflect the higher median CD4 cell count at ART initiation. The gap in CMD prevalence between the urban and the rural setting might also be explained by a high prevalence of adverse living condition in the urban areas: 42% of the HIV positive individuals included in the urban study were unemployed, and 92% reported that they experienced a negative life event (e.g., death in the family, physical or sexual assault, forced eviction, HIV diagnosis, or hospitalisation of the participant or an immediate family member) in the six months before data collection [9,10]. Furthermore, cultural differences in symptoms presentation between urban and rural populations might explain differences in the prevalence between the two settings [33].

Our results confirm earlier data on associations between symptoms of depression and anxiety disorders and suboptimal adherence to ART [12,15]. A meta-analysis of eight studies from low-income countries found that the odds of suboptimal adherence were 92% higher in patients with depressive symptoms than those without depressive symptoms (odds ratio 1.92 95% CI 1.47-2.5) [15]. Another meta-analysis of 11 studies from low-and middle-income countries showed that anxiety disorders were associated with a substantial increase in the odds of suboptimal adherence (odds ratio 1.59, 95% CI 1.29–1.96) [12]. The strength of the association observed in our study was consistent with these meta-analyses.

Our findings show a substantial burden of CMD among people living with HIV in rural Zimbabwe and underline the need to integrate interventions for detecting and addressing CMD in this population. The Friendship Bench project is an evidence-based psychological intervention for delivering mental health care in primary care [21]. The intervention has been rigorously evaluated in a large cluster randomised controlled trial in Harare, which showed that the Friendship Bench intervention substantially improved CMD symptoms [21]. As part of the FB-ART trial, we have implemented the FB intervention at the 8 intervention sites participating in the FB-ART trial, and will further roll out to the 8 control sites to provide access to evidence-based mental health services for people attending these clinics.

Treatment of mental disorders among people living with HIV may also positively affect HIV treatment outcomes [17,18]. A meta-analysis of 29 observational studies and randomised trials showed that depression and psychological distress treatment enhances ART adherence [18]. A further meta-analysis of three randomised controlled trials provides weak evidence for the benefit of cognitive behavioral therapy for depression and adherence on viral load suppression [17,34–36]. Despite these promising results, the evidence for the benefit of mental health care for improving HIV treatment outcomes is still limited.

Further work is needed to evaluate the effect of mental health care on the mental health of people living with HIV and HIV treatment outcomes. We are currently evaluating the effect of the Friendship Bench intervention on ART adherence, viral load suppression and symptoms of CMD among people living with HIV in rural Zimbabwe. There is also a need for continued routine program monitoring and implementation science to ensure the quality and effectiveness of the Friendship Bench intervention in various settings.

In conclusion, our findings show a substantial burden of CMD among people living with HIV in rural Zimbabwe and underline the need to integrate mental health services in HIV treatment programs.

## Data Availability

Data cannot be made available online because of legal and ethical restrictions. To request data, readers may contact IeDEA for consideration by filling out the online form available at https://www.iedea-sa.org/contact-us/

## Ethical considerations

The authors assert that all procedures contributing to this work comply with the ethical standards of the relevant national and institutional committees on human experimentation and with the Helsinki Declaration of 1975, as revised in 2008. The study protocol was approved by the ethics committees of the Medical Research Council of Zimbabwe (MRCZ) (approval number MRCZ/A/2287), the Research Council of Zimbabwe (RCZ), and the Canton of Bern, Switzerland (approval number 2018-00396). Individuals provided verbal consent for eligibility screening and collection and analysis of screening data. Eligible participants provided written informed consent to participate in the randomised controlled trial.

## Authors’ contributions

ME obtained funding for the study. AH, JvD, ME, CK, DC, SH and AL wrote the study protocol. JM and RM assisted with fieldwork and data collection which was overseen by CK and JvD. SH and AH did central data monitoring. AH conducted statistical analysis. AL advised on statistical analysis. AH and wrote the first draft of the paper which was revised by CK, JvD, RV, AL, MH, DC, PvG and ME. All contributed to interpretation of the results, commented on previous versions of the manuscript and read and approved the final manuscript.

